# Identifying a minimal set of single nucleotide polymorphisms to classify the geographic origin of a *P. falciparum* sample from the pf3k database

**DOI:** 10.1101/2022.10.31.22281765

**Authors:** Kyle B. Gustafson, Edward Wenger, Joshua L. Proctor

**Affiliations:** NSWC Carderock Division, West Bethesda, MD 20817; Institute for Disease Modeling, Global Health Division of the Bill and Melinda Gates Foundation, Seattle, WA, USA

## Abstract

Genetic sequencing of malaria parasites has the potential to become an important tool in routine surveillance efforts for the control and eradication of malaria. For example, characterizing the epidemiological connectivity between different populations by assessing the genetic similarity of their parasites can offer insights for national malaria control programs and their strategic allocation of interventions. Despite the increase of whole-genome sequencing of malaria parasites, the development of a small set of single nucleotide polymorphisms (SNPs), often referred to as a barcode, or a panel of amplicons remains programmatically relevant for large-scale, local generation of genetic data. Here, we present an application of a machine-learning method to classify the geographic origin of a sample *and* identify a small set of region-specific SNPs. We demonstrate that this method can automatically identify sets of SNPs which complement the currently targeted loci from the malaria scientific community. More specifically, we find that many of these machine-learned SNPs are near known and well-studied loci such as regions and markers linked to drug resistance, while also identifying new areas of the genome where function is less characterized. The application of this technique can complement current approaches for selecting SNP locations and effectively scales with an increase in sample size.

## 1 Introduction

Malaria genomic surveillance has the potential to provide key insights about the local epidemiology of a region and reveal transmission properties relevant for national malaria control programs (NMCPs). Key programmatic use-cases have been elucidated for malaria genomic surveillance [1], including the identification and characterization of markers connected with treatment failure [2, 3] and the spread of these drug resistance markers [4, 5]. Similarly, parasites with the histidine-rich protein 2/3 deletion evade detection from standard malaria rapid diagnostic tests [6, 7, 8]. Molecular insights such as these have been enabled by technological advances to rapidly and efficiently sequence the malaria parasite genome [9, 10, 11], the curation of global library of malaria parasite genomes [12], and connections to therapeutic studies.

Despite the abundant genomic data and high-throughput deep sequencing platforms, there are still challenges facing the generation of timely, local genetic data. For most high-burden countries, in-country capacity to generate whole genome sequencing data is still limited; relying on out-of-country genomic sequencing centers also introduces operational challenges and delays. Historically, to mitigate these challenges, a smaller set of single nucleotide polymorphism (SNP) positions, often referred to as a barcode, developed with knowledge from a library of whole genomes sequences [9], are utilized to design a rapid, inexpensive assay unique to that setting which can remove delays, build local capacity, and efficiently filter complex genomic data down to operationally relevant parts of the genome. Importantly, these barcodes offer scientists and NMCPs a rapid and tractable path toward scaling genomic surveillance in the near-term. In this article, we demonstrate the utility of a recently developed methodology from machine-learning to optimally select a small number of *region-specific* SNPs that can be used to identify the geographic and temporal origin of a sample.

In addition to identifying and monitoring drug resistant markers, a scale-up of malaria parasite genomic surveillance has the potential to advance current surveillance efforts and intervention design for NMCPs and elimination campaigns [1]. Monitoring disease transmission levels and trends has been linked to features extracted from genomic sequences such as the rate of change of unique versus clonal parasite lineages over time and complexity of infection in a variety of settings [13, 14, 15, 16, 17]. The connection between transmission properties and features from genetic sequences was further strengthened by a modeling study that integrated a mechanistic malaria model with a malaria genome evolution model [18]. Malaria genomic sequences combined with high-fidelity spatiotemporal metadata have also revealed the relative role of parasite movement by characterizing the genetic diversity and gene flow across multiple geographies [19, 20, 5], identifying importation events as compared to local chains of transmission [16, 21], highlighting geographic hot-spots for higher malaria transmission [21, 22, 23], and quantifying connectivity between distinct regions or cities [24]. Each of these use-cases demonstrates how genomic data offers a complementary tool to routine surveillance efforts by NMCPs.

The creation of rapid and inexpensive barcodes based on a small proportion of the whole malaria genome has been integral to investigations on understanding the role of genomic surveillance [13, 14, 15, 25, 26, 18, 27]. Each of these studies use both a different set and number of SNPs, i.e., 5 for Kenya [27], 24 for Senegal [13], approximately 100 for southeast Asia [24], and 384 for Thailand and Malawi [26]. The SNP selection procedures varies widely across studies including, but not limited to, picking SNPs or regions of the genome with a high minor allele frequency [13, 24], a large proportion of total genetic variance in a region represented by a specific geographic sub-population [28], the relationship between expected heterozygosity and within-Africa genetic differentiation [29], backward elimination locus selection and cross validation [30, 27], and repeating analyses with ever decreasing numbers of random subsets until estimates become unreliable [24].

Here, we adapt a recently developed methodology, called sparse sensor placement for optimal classification (SSPOC) [31], to identify the minimal number of SNPs required to identify the geographic origin of a sample. SSPOC is fundamentally data-driven; it uses publicly available genomic data, the global pf3k project, to select the minimal amount of information required to perform a classification task. It has been applied to a variety of machine-learning tasks including facial recognition and sensor placement in a fluid flow field [31]. The SSPOC methodology offers a number of advantageous characteristics for this investigation: the method is uniquely suited for classification problems with many more measurement elements than samples, which is often found with genomic data; SSPOC is also interpretable selecting a small number of *specific* SNPs; the SNPs are chosen to be support the discrimination task based on the geometric structure of the available data and *not* along principal component directions; the optimization process is convex, enabling efficient numerical solver to rapidly converge to a solution; and the method leverages a number of recent, foundational results in statistics and machine-learning [32, 31]. We demonstrate how SSPOC can be used to efficiently generate a setting-specific barcode, quantify the number of SNPs required to achieve a classification accuracy close to to using all genetic positions, and compare to random subsets.

## 2 Study data and methods

### 2.1 Malaria genomic surveillance data

We utilize malaria parasite genomic data collected in the Pf3k project from the Malaria Genomic Epidemiology Network [33] which contains samples from 26 sites across Africa and Southeast Asia. A total of 2076 samples with sequenced reads at nearly 39452 single nucleotide polymorphisms (SNPs) were obtained for both the reference and alternative allele sequencing depth. Each genomic sample is associated with metadata which includes the sampling site and, for a subset of samples, the date of collection. The samples were nearly evenly split between Africa and Asia (1099:977). Seven African countries were represented. Two sites contributed a large majority of samples from the continent: Ghana (478) and Malawi (262). Seven Southeast Asian countries were represented by sampling, including the Ramu site in eastern Bangladesh near the border with Myanmar (Burma), at the edge of the Southeast Asian region. Over half of these samples came from four data collection sites throughout Cambodia; see Fig. 1A for an illustration.

**Figure 1:**
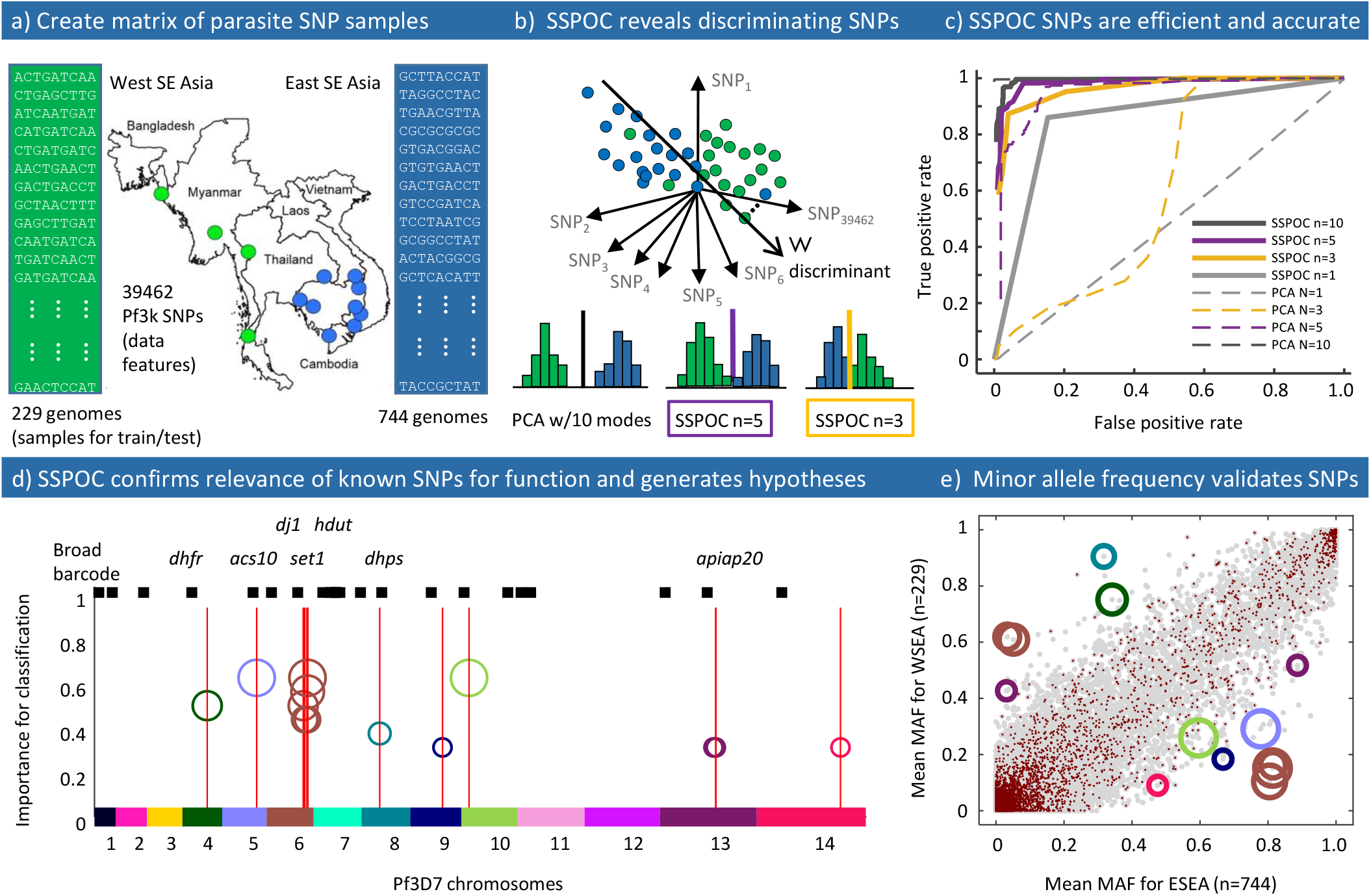
Summary of data and methods shown with highlight from results: a) A map of collection sites in Eastern and Western Southeast Asia is combined with the concept of a data matrix for machine learning on genomes. b) The dimensionality of genomic SNP data is very high, but the sparsity of distinguishing SNP alleles reduces the complexity of the classification problem. c) The true positive rate compared to the false positive rate (ROC curve) for discrimination between Eastern and Western SE Asia is shown, with several examples colorized to match the scheme in panel b). Most notably, a small number of SSPOC SNPs performs better than the same number of PCA modes and provides interpretable genomic loci. d) The genomic loci of SSPOC SNPs shows both agreement with known drug resistance loci and new genomic loci for further study. Red vertical lines indicate named genes with an intergenic, non-synonymous SSPOC SNP with frequency*>* 30%. The size of the circles that mark the SSPOC SNPs are scaled by frequency of appearance in multiple test/train splits and match the circles in panel e). The black barcode above the red lines indicates the loci Harvard/Broad barcode SNPs. e) MAF bias scatter plot for WSEA and ESEA where each gray point represents one SNP, averaged over all the samples in each category. SSPOC SNP loci are indicated with circles sized by the same scale in panel d. Red points are SNPs that have more than the overall average number of sequencing errors across the samples in either category, which are *a priori* excluded to prevent bias.

### 2.2 Known *P. falciparum* SNPs

We collected a number of popular and functionally relevant SNPs from previous studies, including the first malaria barcode of 24 SNP positions [13]. We manually compiled a list of 23 SNPs with large non-reference allele frequency (NRAF) from the MalariaGEN consortium Panoptes portal [12]. In addition, we have also included *kelch13* SNPs which have been associated with Artemisinin-resistance in the Greater Mekong subregion of Southeast Asia [33]. See Fig. 1D for a depiction of the location of these SNPs along the malaria parasite genome.

### 2.3 Transforming major and minor allele reads to a categorical variable

Deep sequenced samples in the pf3k project provides a read depth for both reference (major) and alternate (minor) alleles at the SNP positions described in §2.1. The minor allele frequency (MAF), computed by the number of minor alleles divided by the total read depth (minor and major alleles), at each SNP creates a fingerprint for each sample. For this work, We encode this fingerprint into a categorical variable at each SNP position. Each SNP for each genome was assigned a category, (*a, b, c*), where *a* = *b* = *c* = 0 unless: *a* = 1 if the allele is reference, *b* = 1 if the allele is alternate, and *c* = 1 if a mixed position is indicated. In order to be categorized as a reference or alternate call, the number of alleles measured at that position must be greater or equal to five *and* the opposite allele must be less than five reads. This account for potential measurement error at each position and only accounts for less than 1.2% of positions. A mixed position category indicates that there is a read depth of both minor and major allele above five, occurring at a rate of approximately 4.4%. The resulting data matrix, **X**, contains the three digit category key for each of the 39452 alleles measured in each of the 2076 parasite genomes, resulting in expanded dimensions of 118356 *×* 2076. Note that we could have used a two category key for this expansion, however, in future analyses we aim to expand this approach into loci beyond solely bi-allelic SNPs.

### 2.4 Reducing dimensionality via the singular value decomposition

The singular value decomposition (SVD) is a matrix factorization technique that has been extensively utilized to reduce the dimensionality of datasets that contain many more dimensions than number of samples in order to perform standard statistical analyses [34, 32]. The SVD algorithm is at the root of well-known and mathematically related methodologies such as Principal Component Analysis (PCA) [35], the Hotelling transform [36], Empirical Orthogonal Functions [37, 38], Proper Orthogonal Decomposition [39], and the Karhunen-Loève decomposition [40]. The SVD of the genome feature matrix **X**, defined in §2.3, is given by **X** = **ΨSV**^*\**^, where *n* = 118356, *m* = 2076, **Ψ** ∈ ℝ^*n×n*^, **∑** ∈ ℝ^*n×m*^, **V** ∈ ℝ^*m×m*^, and *\** denotes the complex conjugate. We consider a standard factorization which sorts the singular values of **∑** by decreasing magnitude. The dimensionality of **X** can be reduced from *n* = 118356 to *r* utilizing the first *r* left singular vectors, denoted by 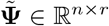, and the linear projection 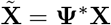. Thus, each genome representation of **x** ∈ ℝ^*n×*1^ is transformed and reduced to 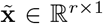. The Eckart-Young theorem [41] provides the theoretical foundation for taking the first *r* singular vectors and singular values as the best rank *r* approximation of **X**. Unless otherwise stated in the text, we use *r* = 40.

### 2.5 Predicting classes with linear discriminant analysis

The statistics and machine-learning (ML) literature offers a wide-variety of supervised discrimination algorithms to predict class origin. Here, we utilize linear discriminant analysis (LDA) as a parsimonious and interpretable algorithm to discriminate the geographic origin of a sample within the malaria genomic data. Other ML algorithms may provide slightly better accuracy, however, the linearity of LDA is integral to the sparse optimization described in the next section. LDA reveals a single discrimination vector, **w**, that optimally separates classes of data points by solving the following eigenvalue problem. The vector **w** is computed by:

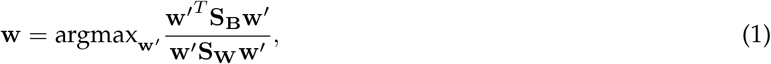

where **S**_**B**_ is the between-class scatter matrix and **S**_**W**_ is the within-class scatter matrix. These matrices are computed as: 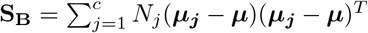 and 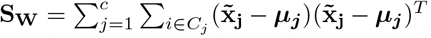, where *c* is the number of classes, *N*_*j*_ is the number of samples in class *j, C*_*j*_ is the set of samples in class *j*, ***μ*** is mean of all samples, and ***μ***_***j***_ is the mean of the samples in class *j*. Once the discrimination vector has been computed, a threshold value assigns the sample to a class, such as “Eastern SE Asia” and “Western SE Asia.” This is also the parameter that governs the balance between sensitivity and specificity for the discrimination task; see §2.7 for more details. Figure 1(B) shows three illustrations of a discrimination vector, projection of data samples on to the vector, and a threshold for defining class assignment.

### 2.6 Sparse sensor placement for optimal classification (SSPOC) for malaria parasite genomes

SSPOC is a recently developed algorithm based on *l*_1_ minimization that identifies a small number of individual elements, in this case SNPs, that discriminates between classes [31]. SSPOC solves for the measurement vector of SNPs **s**, which will have a maximum of *r* nonzero entries:

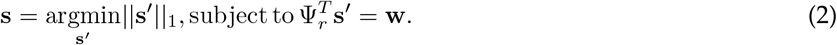

Here, **w** is the discrimination vector in feature space Ψ_*r*_, which can be an LDA vector in a PCA space as described above. We tested two types of sparsity-promoting algorithms for solving this core data-driven *l*_1_ minimization problem, as described in the Supplement.

As shown previously [31], the sparse set **s** of measurements can be interpreted as the sum of dominant and non-dominant features for discriminating between classes. A major advantage of using **s** rather than the PCA solution is that the number of measurements can be further reduced by comparing classification accuracy either to all SNP data (PCA) or to self-convergence with number of SNPs. We refer to such a minimal set of SNPs as SSPOC SNPs. Note that the linear discriminant analysis can be efficiently expanded beyond a two class discrimination task.

### 2.7 Evaluating accuracy of genome classifications

Classification tasks, whether through ML or human subject matter expertise, are subject to errors that can be minimized with appropriate data collection and more finely tuned models [42]. Here we are mainly concerned with the binary classification of genomes into one of two geographic origins, A or B, that are assumed to contain all of the samples. We used the SNP categories as features for input into a standard PCA/LDA classification task using an 80*/*20%, test/train split from the available genomes of known origin by sampling site. Multiclass discrimination across several origins can be viewed as a generalization of binary discrimination when defining class A in contrast to all other classes. The linear, two-class classification approach can be quantified and visualized with a receiver operator characteristic (ROC) curve, which is a plot of true positive rate (detection) against false positive rate (false alarm) as a discrimination variable is varied. For determining geographical region of origin by SNPs, we computed the ROC curve by varying the threshold on the **w** where a sample is assigned to a class A or B. We computed ROC curves for all data (PCA), sets of SSPOC SNPs of various size, and randomly chosen SNPs. As shown in Fig. 1C, the behavior of the ROC improves to a limiting curve as the number of SSPOC SNPs increases from *N*_*SNP*_ = 3 to *N*_*SNP*_ = 20, for the case of Eastern/Western SE Asia. We also define the converged error rate of a classification algorithm as the fraction of false negatives remaining when the number of measurement (SNP) locations used is very large. The converged error rate when using all the SNPs (PCA) sets a lower bound on the classification error. This lower bound can be large (approaching 50%) when the sampling sites are near to each other and small (less than 5%) when the sampling sites are well-separated.

### 2.8 Data Availability

All data used in this article is publicly available [33].

## 3 Results

### 3.1 Baseline classification accuracy achieved with PCA and LDA

A baseline for evaluating the success of selecting a subset of SNPs using SSPOC is to leverage all available SNPs using PCA and LDA. For example, the converged validation error is 2% for genome discrimination between sampling sites in Guinea (100 genomes) and DRC (107 genomes) that are well separated by distance and geographic barriers. Classification accuracy using 40 PCA modes is higher compared to a randomly chosen set of 40 SNPs.

The same analysis shows a similar error level of less than 5% between genomes known to be from East SE Asia (744 genomes) and West SE Asia (210 genomes); see Figure 1C. for the ROC curve. In contrast for Guinea (100 genomes) and its neighbor Mali (84 genomes), the baseline classification error using the standard PCA/LDA method is 20%. This larger error compared to the well-separated sampling regions is consistent with the expected correlation between geographic proximity and genetic similarity.

### 3.2 SSPOC identifies a parsimonious set of SNPs for highly accurate classification

SSPOC can achieve high classification accuracy by leveraging a small number of SNPs and their characteristics to discriminate between specific populations of parasite lineages. Figure 1C shows the convergence of ROC curves for a range of SSPOC SNP counts(see Sec 2.7) for the West/East SE Asia classification task. Also shown in Fig. 1C is the ROC curve for the baseline PCA method with 40 singular vectors using all SNP data. The ROC curve for SSPOC calculated with *N*_*SNP*_ *≈* 10 very quickly reaches *TPR >* 90% for *FPR <* 10%, which is qualitatively similar to the PCA baseline shown on the same axes. An equally small number of randomly selected loci give a converged error rate near an upper bound of 50%.

### 3.3 SSPOC SNPs are interpretable and often consistent with known functional loci

In addition to the improvement in classification accuracy with fewer SNPs, SSPOC SNPs point directly to the genomic positions rather than the weighted average of multiple positions given by PCA. Many of these SSPOC SNPs are located near known anti-malarial resistance loci and SNP locations previously identified in the Broad/Harvard barcode (*pfdhfr* [43], *pfcrt*[44], *pfnif4* [45]). Other SSPOC SNPs are proximal to genes with known functions beyond anti-malaria resistant markers (*pfgarp* [46], *pfset1* [47, 48]). This is shown in Fig. 1D, where the number of occurrences of a SSPOC SNP over several test and train iterations of the LDA classifier for Eastern/Western SE Asia (WSEA/ESEA) are shown across the *falciparum* genome, color-coded by chromosome number (binned at a resolution of 1 kb). These SSPOC SNPs are plotted as open circles at a height and with an area proportional to the number of times they are selected across ten SSPOC cross-validations.

In Chr 4, the *pfdhfr* SNP is exactly in the center of the *≈* 2kb gene. Three other SSPOC SNPs are intergenic to *pfdhps* (Chr 8), *pfapiap2* (Chr 13), and *pfset1* (Chr 6) proximal SSPOC SNPs. Seven other SSPOC SNPs are within 100 kb of the center of important genes (*pfmdr1, pfcrt, pfwd102, pfdhfs*). Figure 1E shows minor allele frequency (MAF) for the same SSPOC SNPs using the same color code for WSEA/ESEA. The correlation between high MAF values and SSPOC SNPs highlight the consistency of the method with other standard approaches, but also shows where there are differences. We also found six SSPOC SNPs within 100 kb of Broad/Harvard barcode SNPs on Chr 1, 2, 7, and 8, as shown in Fig. 1D.

### 3.4 Distance between sampling sites predicts the number of SNPs to classify samples

We found a clear trend of decreasing sample classification error with the distance between sampling sites. Figure 2a shows LDA classification error compared to physical site separation using the first 40 modes from PCA, which was found to be a sufficient number of features for the error to converge. Here, the separation distance is taken to be the driving distance computed using the Google Maps API Glocalsearch to provide a common metric for the diverse sample of sites. Driving distance is a proxy for effective mixing between populations, which neglects cultural or economic connections that are beyond the scope of this work to measure. The minimal error decreases to essentially zero within 2000 km for SE Asia, but is not observed to be zero until 4000 km for Africa, partly due to fewer sampling sites available to compare. All intercontinental site pairs show zero error when using the first 40 PCA modes. Sparse sets of SSPOC SNPs exist for all national and subnational pairs of sampling sites, as shown in Fig. 2b. When defining a match between the SSPOC and PCA error as less than 5%, it is sufficient to use fewer than 42 SNPs for any pair of sites. A decrease with distance in the number of SNPs necessary to discriminate between origin sites is observed generally, though there are several anomalies for both Africa and SE Asia. Often, far fewer than 40 SNPs are required to acheive the baseline accuracy, even for some nearby sampling sites. One exceptional case, for Ghana compared to Mali (see also Fig. 12), is due partly to the presence of multiple SNPs in *pfdhps, pfdhfr*, and *pfcg2*. For this example, it is possible to achieve 10% better accuracy compared to the baseline 40 PCA modes. These results provide direct quantification of the correlation between physical distance and genomic similarity while highlighting the significance of a small number of SNPs for efficient geographic classification in each case.

**Figure 2:**
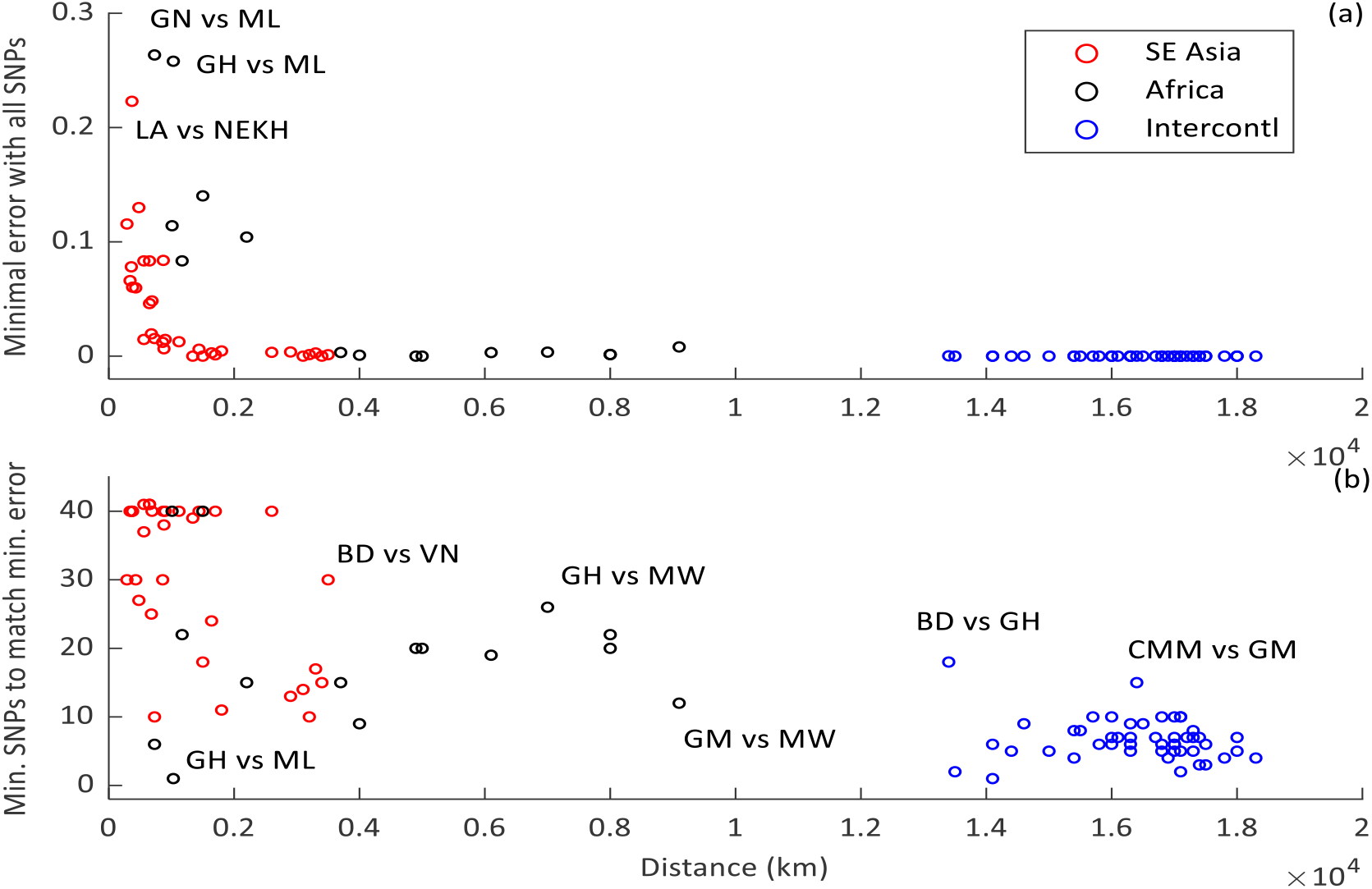
These two panels show the travel distance-dependent trends in the minimum geographic classification error and the number of SSPOC SNPs required to converge to that error. In panel (a) the minimum binary classification error achievable using linear discriminant analysis (LDA) for 40 principal component (PCA) modes is shown for several pairs of sample collection areas within and between Africa and SE Asia as a function of the driving distance between sites. Decreasing error is observed for Africa (black circles) and Asia (red circles), while for the intercontinental pairs, zero error is always approachable. The decrease with distance for SE Asia is better characterized due to a larger number of sampling sites at short distances, though it does appear that the error drops faster with distance for SE Asia. Panel (b) shows the number of SSPOC SNPs, calculated up to 42 SNPs, required to approach the PCA results within *<* 5%, as a function of driving distance. In many cases, 40 SNPs are required to achieve the 5% margin, though there are many examples where far fewer SSPOC SNPs can be used. For the intercontinental pairs where a larger number of SSPOC SNPs are required (CMM/GM, BD/GH), SSPOC has been affected by small sample sizes. Error, SSPOC SNP counts, and driving distance for each pair are available in the Supplemental Files.

### 3.5 SSPOC expands candidates for SNPs to be added to barcodes

Two detailed examples of pair-wise classification are shown in Fig. 3: Laos versus Vietnam (Fig. 3a-c) and Ghana versus Guinea (Fig. 3d-f). For both comparisons, the importance of a SNP for classification is indicated by the fraction of occurrences (vertical axis in Fig. 3a,b,d,e) of the SNP across a range of test sets that vary both across a random subset of the data and the allowed number of SNPs for the comparison (from the MAF-rank list and using the SSPOC sparsity parameter). The scatter plots (Fig. 3c and f) display the site-averaged MAF for all the SNPs with grey dots, highlighting the selected SNPs for both MAF-rank (cross symbol) and SSPOC (plus symbol) in terms of classification accuracy using the same color scheme as for the chromsome maps. These plots represent one illustration of the results when limiting the classification test splits to contain *N* ^*\**^ + */ −* 2 SNPs, where *N* ^*\**^ is number of SNPs required to approach the 40-mode PCA classification accuracy within 5%. Both of these plots also display only SNPs that appear in more than 20% of the classification sets within the *N* ^*\**^ + */ −* 2 range. More detail on these examples, including the accuracy of the classification, is shown in Fig. 10 and Fig. 11 and included in Supplemental Data.

**Figure 3:**
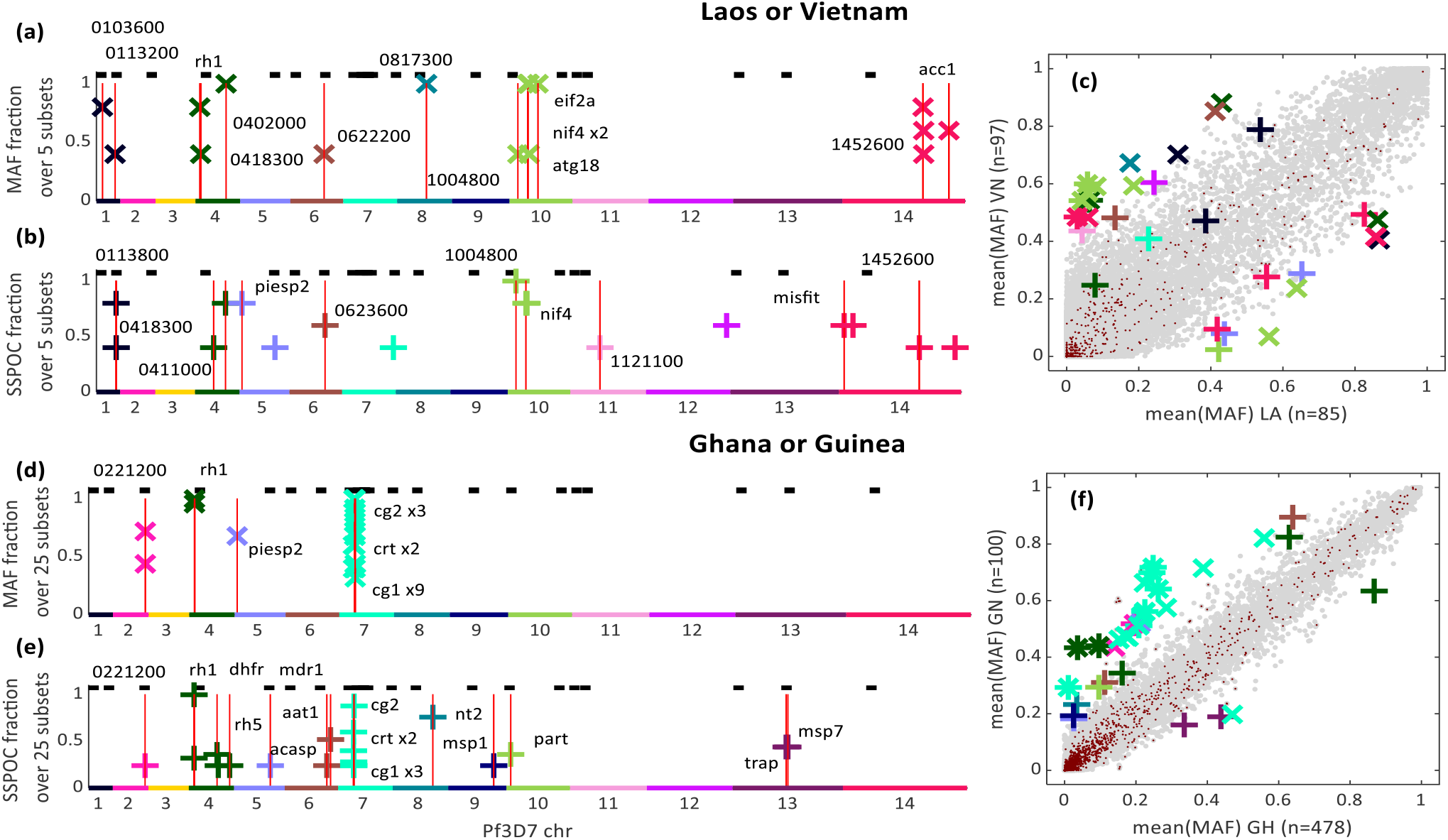
Two examples of binary national level geoclassification using MAF-rank and SSPOC SNPs. Red vertical lines indicate named genes with an intergenic, non-synonymous SSPOC SNP with frequency*>* 30%. The black barcode above the red lines indicates the loci Harvard/Broad barcode SNPs. Gene names (PlasmoDB) are displayed for a subset of SNPs for illustrative purposes, while the full list of SNPs is in the Supplement. Scatter plots of MAF, averaged over genomes for each site, for the entire set of Pf3k SNPs (grey dots) are shown. The the number of genomes for each nation is shown on the axes labels, while the chromosome color coded SNPs are highlighted with ‘x’ for MAF-ranked and ‘+’ for SSPOC. SNPs with sequencing errors occuring more frequently than one standard deviation above the population mean are excluded from the analysis and indicated with red dots. In a)-c), Laos and Vietnam genomes are classified, while d)-f) shows SNPs important for classification of Ghana versus Guinea.

There are many genes in which non-synonymous SNPs appear with high frequency across the tests and also within which several SNPs are clustered. For the neighbors in SE Asia (LA vs VN) the MAF-rank method (Fig. 3a) indicates *pfnif4, pfatg18*, and *pfeif2a* as well as two unnamed genes (*pf3d70418300* and *pf3d70817300*) are relevant for classification. The same data classified with the SSPOC method (Fig. 3b) shows some agreement (*pfnif4* and *pf3d704180300*) but also suggests the importance of other SNPs such as *pfpiesp2*[49, 50], *pfmisfit, pf3d70113800*, and *pf3d70623600*. The scatter plot (Fig. 3c) reiterates that there is little exact agreement on SNPs between MAF-rank and SSPOC, except on *pfnif4, pf3d71004800*, and *pf3d71452600*. Among these genes, NLI Interacting factor-like phosphatase (*pfnif4*) is known to be relevant to drug resistance.

Genomes in the West African nations (Ghana vs Guinea) are separated by a wider variety of SNPs, potentially indicating greater internal diversity of genomes. Nevertheless, both MAF-rank (Fig. **??**d) and SSPOC (Fig. **??**e) show agreement on some of the most discriminatory SNPs including *pfcg1, pfcg2, pfacasp, pfcrt, pfrh1* and *pfdhfr*. Previous GWAS[51] and functional[52] studies have shown the importance of *pfcg1* and *pfcg2* to chloroquine resistance, though this is attributed to linkage disequilibrium with*pfcrt*. At this level, SSPOC uniquely selects for drug resistance genes, with two SNPs in *pfmdr1*, one in *pfmdr2*, as well as one in *pfnt2*[53] and three more in *pf3d70710200*. Additionally, SSPOC finds multiple SNPs separating Ghana and Guinea in two merozoite surface protein genes (*msp1* and *msp7*) that are known to be required for host cell invasion[54].

### 3.6 Generalization of SSPOC to multiple geographic categories

We also performed a multisite, global classification between Laos, NE Cambodia, N Cambodia, Vietnam, W Cambodia, and Thailand (see Fig. 4). This classification task was better than 90% successful for four of the six classes using 50 SSPOC SNPs, with Laos and Vietnam performing at 60% and 70% respectively. See Supplemental Data for more detail on the convergence of performance metrics for this classification. Using this set of SNPs, all of the misclassifications of Laos and most of Vietnam were to NE Cambodia. The most frequently observed SNPs is an annotated gene of unknown function: *pf0216500*. The next most frequently occuring SSPOC SNPs across the five test splits include genes with known functions *pfrcc1*[55], *pftrap*[56, 57] *pfnif4, pfulg8*[58], and *pfercc4*[59]. Note that the misclassification of samples represented in Fig. 4 may not be in reality a misclassification, but instead a measure of connectivity or importation between regions; this will require further study and validation.

**Figure 4:**
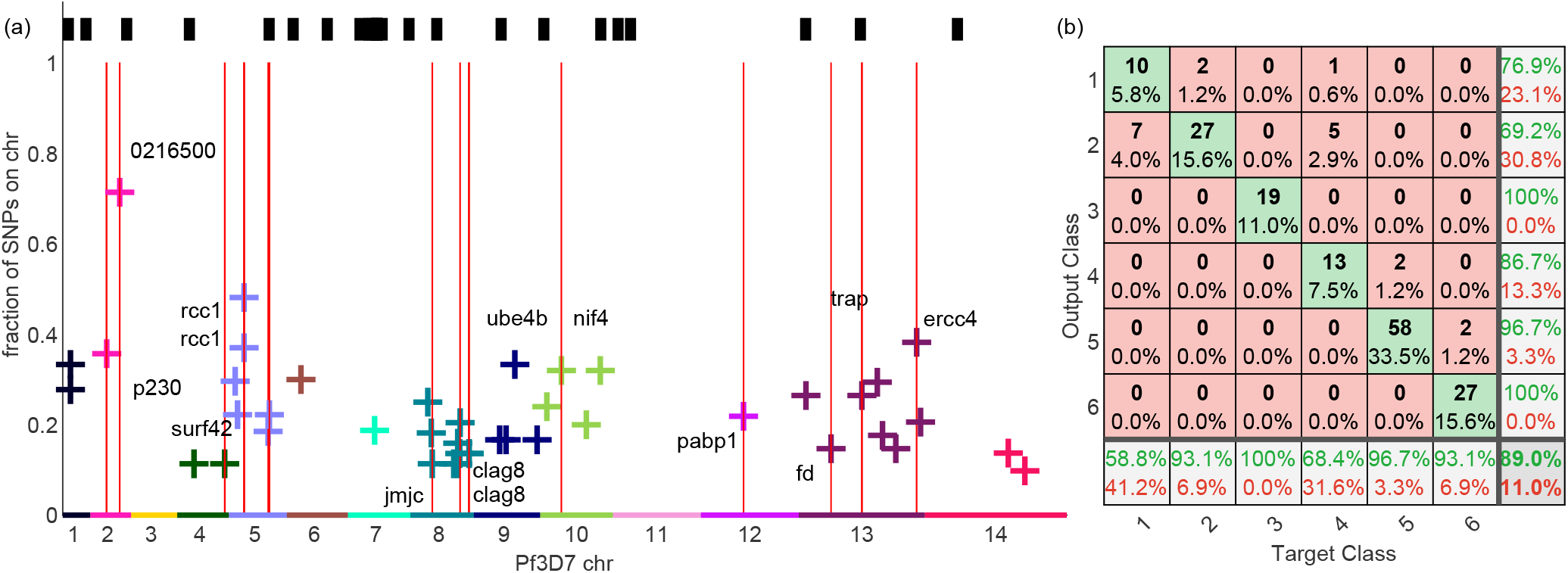
Multiclass discrimination by SSPOC SNPs for six subregions in SE Asia, including three subnational areas of Cambodia defined by testing site locations. These areas are Laos, NE Cambodia, N Cambodia, Vietnam, W Cambodia, and Thailand. For these panels, five test/train splits of 80/20 were used to find the most informative SSPOC SNPs and compute the average classification accuracy. In (a), the locations of SSPOC SNPs that appear most often for the six class task are plotted by their frequency of selection by SSPOC, indicating notable genes by name. Red vertical lines indicate named genes with an intergenic, non-synonymous SSPOC SNP with frequency*>* 30%. The black barcode above the red lines indicates the loci Harvard/Broad barcode SNPs. In (b), a confusion matrix is shown for 50 SSPOC SNPs averaged over the five test splits, listed in the same order as above. The central cells of the confusion matrix contain both the number of samples and percentage of the total samples from the test sets. Each of the five test sets contains 173 samples, so that the 58 samples from W. Cambodia that were correctly identified by SSPOC represent 33.5% of the total samples. For each SSPOC predicted output class, the far right column in (b) shows the precision (green) and false discovery rate (red), while the bottom row shows the true positive (green) and false negative (red) rates. The green percentage in the lower right, 89%, is the overall accuracy for the multiclass model with 50 SSPOC SNPs.

**Figure 5:**
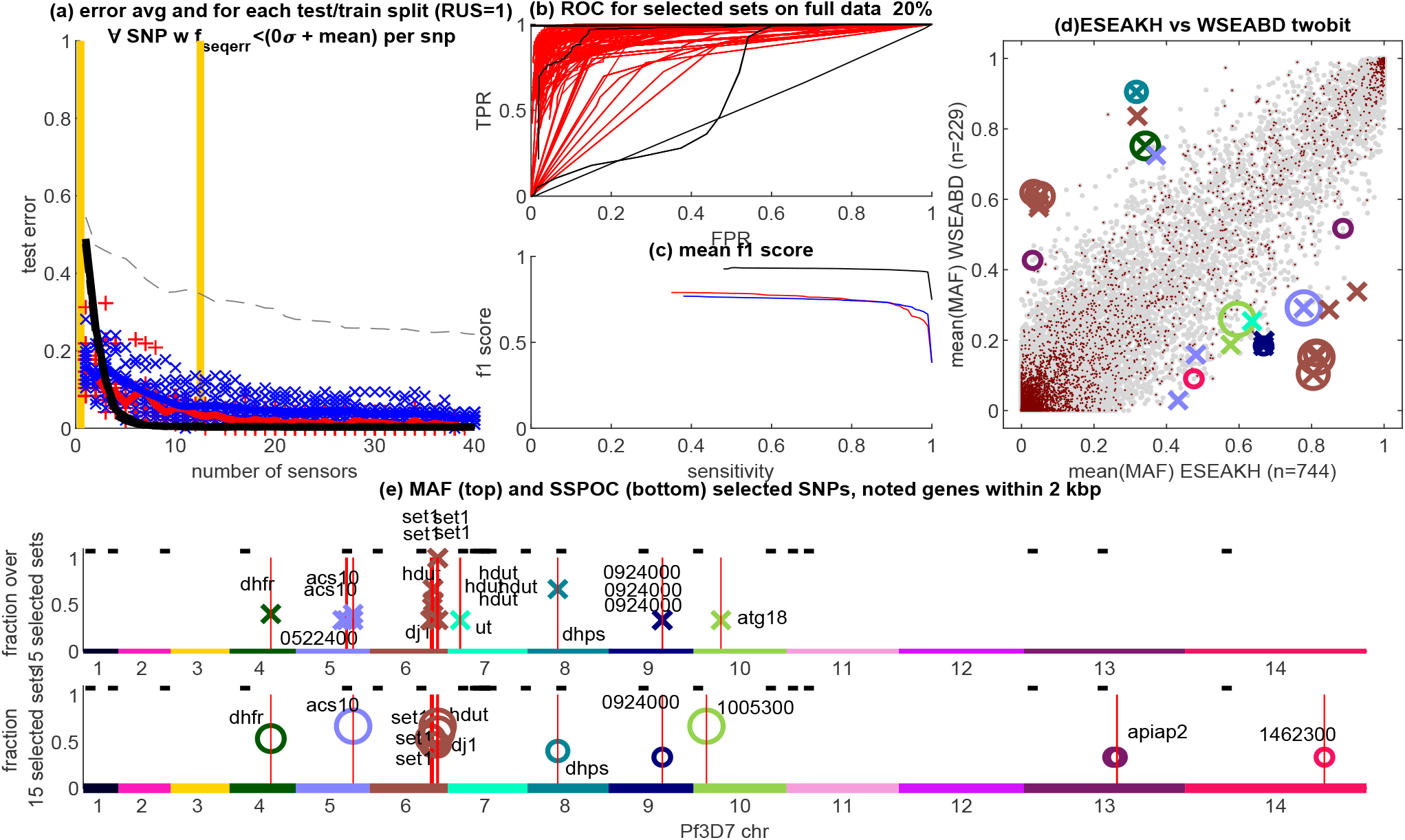
Expanded view of data analyzed for Figure 1. The error rate for PCA, SSPOC, and MAF ranking is shown in a), while ROC and f1-score is shown in b) and c). Panel e) shows the genomic positions for the highest ranked MAF-biased SNPs and the SSPOC SNPs from the groups selected in panel a) between the yellow lines.

### 3.7 Seasonal differences in parasite populations may be detectable with SSPOC SNPs

In addition to improving the efficiency of determining the geographic origin of a *P. falciparum* genome, the SSPOC method should be useful for determining the season of the sample collection. In this dataset, most season to season collections are too imbalanced to inform any classification method. Seasonal collection was undertaken in two countries: Malawi and Ghana, as shown in Fig. 6. As a test case for seasonal classification, we examined the seasonality of genomes in Chikwawa, Malawi between 2012 (81 genomes) and 2014 (25 genomes). These genomes were used to train a SSPOC classifier using both the correct season and a random permutation of the seasonal label as a negative control.

**Figure 6:**
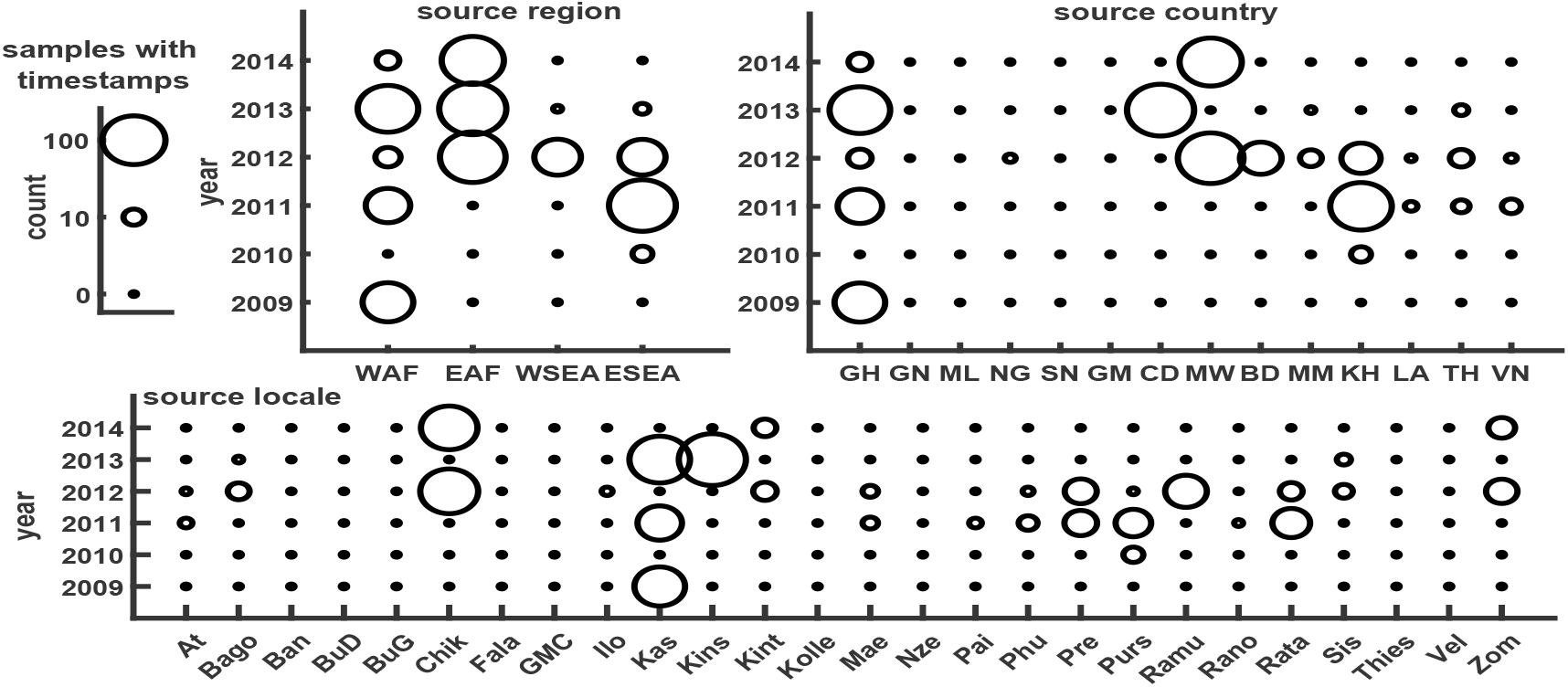
Visualization of counts of available parasite genomes categorized by year and location of collection. The size of the circle scales with the number of genomes in the spatiotemporal bin indicated by the axes of the plots. The missing samples shown here compared to the previous table are due to many samples missing a collection date in the available metadata. Three plots show the spatial location at three scales: regional, national, and sampling site locale. Nationally, a strong sampling bias exists towards parasites collected from Cambodia, Ghana, and Malawi. Nigeria has only five samples, while most other countries have more than 50 genomes. Temporally, even on the large-scale regional level, there are large gaps from year to year. All of the East Africa samples are between 2012-14, while most of the eastern Southeast Asia samples are from 2011.

SSPOC converged accuracy for these genomes is relatively low for both the correct and the scrambled labels, compared to the well-separated geographic results presented previously. However, as shown in Fig. 18, the genomic locations of the SSPOC SNPs for 2012-14 are more coincident with the known locations of resistance genes. Moreover, MAF values for the randomly permutated labels are much smaller than for the actual temporal classification and the MAF ratio is much closer to parity than for geographic separation.

## 4 Discussion

Malaria genomic surveillance is gaining momentum toward integration into routine surveillance activities and could be an essential operational tool for national malaria control programs over the next decade. However, the integration of genomic surveillance is constrained by several factors including limited local capacity for wholegenome sequencing in malaria endemic countries, operational challenges around using a small number of high throughput sequencing centers, and knowledge gaps around several key malaria genomic use-cases beyond the spread of drug resistance and diagnostic failure phenotypes. Mitigating these challenges in the near term will require judicious selection of samples for whole-genome deep sequencing and the development of region-specific barcodes or amplicon panels that can be generated at scale within malaria endemic countries. For this approach, a key challenge is the selection of genomic regions that provide information for malaria genomic surveillance usecases. In this work, we implement a new approach designed to identify a small number of individual SNPs from a library of whole genome sequences that can be used to discriminate between geographic populations. We demonstrate the framework on genomic sequences available in the pf3k project and show the results are both parsimonious and interpretable allowing comparison to the broader malaria genomic literature. This approach can be efficiently scaled up by number of samples and geographies for new data collection efforts.

Our results are broadly consistent with previous studies. We find a similar number of SNPs are required at different geographic distances. For example, we have very good accuracy of classifying the origin of a malaria sample between Africa and Southeast Asia continent requiring only a few SNPs (*≈* 5); this has been previously noted when randomly subselecting SNPs in pf3k. Restricting the geographic regions to the country scale, we find the number of SNPs required to be on the order of 20 *−* 30, matching several previous studies focused at the country or multi-country scale [13, 26, 27]. We also find that several SSPOC identified SNPs are in regions near well-known SNPs from the literature including the *kelch13* loci and overlap with SNPs identified using a ranked minor allele frequency approach. Both the number and location of the SSPOC SNPs are well aligned with the literature.

Our framework and results have also broadened the scope of previous investigations. We have identified the number of SNPs required to classify the origin of a sample for a wide variety of setting comparisons and quantified the predictive power of these SNPs. Moreover, we have demonstrated how the accuracy of our methodology converges to an upper limit, where we use all available positions, as compared to randomly grabbing a number of SNP positions from the genome. For some cases, this difference is substantial, i.e., *≈* 80 positions; see §6 for more details. Brunton et al. [31] previously noted that the SSPOC algorithm outperforms random measurements suggesting the methodology recovers SNPs with more information content. Importantly, this method also highlights regions of the genome that have not, to the author’s knowledge, been previously documented or identified. In addition to providing a setting-specific set of SNPs for an assay, this could also be a tool to identify functionally important positions for investigation. Combining this approach with others from the literature would provide practitioners a more complete set of candidate SNPs for their barcode or amplicon development.

There are several key challenges facing this approach. Similar to other methods, the algorithm requires a library of parasite genomic sequences from a particular geographic setting. Here, we have used the global open database pf3k [12], but despite the growing number of available sequences in projects, such as the recently released pf7k [28], these samples are likely not representative of every malaria endemic region, nor genetic population subgroups within those regions. An upfront cost will be required for WGS of samples in new regions as well as a re-sampling scheme using WGS as the underlying genetic population may change due to drug and intervention pressure over time as seen with drug resistance makers in the Greater Mekong Subregion [60]. Additionally, we have framed our approach in terms of identifying single SNPs by encoding the value of that SNP in terms of a biallelic position that can either be the reference, alternate, or mixed alleles as well as too low sequencing depth. Extensions of the approach to multi-allelic loci is quite natural, however, extending this approach to explicitly account for microhaplotype structure for a panel of amplicons is possible, but will require methodological development. We also assume that the sample collection location of the malaria parasite is the origin of the infection; this may not actually be the case due to human and parasite movements, thus potentially affecting the SNP selection process.

Despite these challenges, our approach is quite timely given the scale-up of genomic surveillance and sequencing of malaria parasites across sub-Saharan Africa. The combination of WGS, barcodes, and amplicon panels will continue to be practical in the near-term. Methodological approaches, like the one presented here, can complement other approaches and support malaria genomic use-cases such as identifying population connectivity and the relative role of importation. The combination of new data generation and development of innovative molecular and methodological tools are key near-term investments for the longer term global goal of malaria eradication.

## Data Availability

All data in this article is publicly available [33].

## 5 Acknowledgments

The authors would like to thank Samir Bhatt and Ewan Cameron for productive early conversations on this research problem and approach. K. B. G was funded in part by the Naval Innovative Science and Engineering (NISE) program, managed by the NSWC Carderock Division Chief Technology Office. K. B. G. performed analyses and wrote the paper. E. W. devised methods and revised the paper. J. L. P. designed the methods, directed the research, and wrote the paper.

## 6 Supplement

### Supplemental Information

We have made the source code and data files for our analysis available at github.com/kgustafIDM/.

#### 6.1 Spatiotemporal distribution of collection sites

The compilation of genomic studies by the Pf3k consortium (https://www.malariagen.net/projects/Pf3k) produced a large dataset across parasite collection sites in Africa and Southeast Asia. Table1 shows the number of genomes sequences from each country of collection in this group of studies. In Figure 6, we also visualize the count of genomes as they are separated by year and location of origin at several levels of geographic detail: region of the globe, country, and locale of the collection site village. It is clear that some of the collections sites have far more samples than others, while some years are highly over-represented in the full data set. Therefore, the geographic classification results primarily highlighted in this work are based on combining the genomes collected each year from 2009-2014 for a given location.

**Table 1:** Summary counts of *P. falciparum* genomes from the Malaria Genomic Epidemiology Network used in this study, sorted by geographic origin.

| Country | Region | Number of Pf3k genomes |
| --- | --- | --- |
| Bangladesh | West SE Asia | 48 |
| Myanmar | West SE Asia | 58 |
| West Thailand | West SE Asia | 104 |
| Laos | East SE Asia | 85 |
| Vietnam | East SE Asia | 97 |
| East Thailand | East SE Asia | 21 |
| Cambodia | East SE Asia | 541 |
| Guinea | West Africa | 100 |
| Mali | West Africa | 84 |
| Nigeria | West Africa | 5 |
| The Gambia | West Africa | 63 |
| Ghana | West Africa | 478 |
| Malawi | East Africa | 262 |
| D.R. Congo | Central Africa | 107 |

#### 6.2 Efficiency of numerical solvers for sparsity in SSPOC

We compared the stability and speed of two methods, SPGL and CVX, for solving the central numerical algorithms in SSPOC, which is the sparsity-promoting step. SPGL is a faster, first-order matrix-based method for solving one-norm regularized least squares. CVX is a more general framework for convex optimization, which was implemented in this analysis with a relatively slow predictor-corrector method for determining the maximally sparse solution. Both numerical methods show similar results up to 100 SNP positions tested when averaged over 10 test/train repetitions. CVX is more stable for larger numbers of sensor locations, whereas the theoretically ideal SPGL solution is inherently more difficult to find due to a large number of similarly optimal values. This property of the solution causes SPGL to behave erratically for larger numbers of SNPs, though the *>* 10*x* speed benefit may be preferred for some applications, such as for very large datasets or parametric analysis using specific subregions of the genome.

#### 6.3 Detailed results for 102 pairs of sampling areas at national or subnational resolution

Table S2 (Excel file) lists the driving distance between each pair of national level (and some subnational) sites along with the PCA (40 mode) error in binary classification and the number of SNPs needed to approach that error within 5% for both SSPOC and MAF-rank methods. Here we provide tables (Table S3a and Table S3b as Excel files) for the SNPs that are selected by SSPOC and MAF-rank that appear in the classiification test sets when the number of SNPs per set falls within a range within *N \** +*/ −* 2 of the number of SNPs listed in Table S2. Table S3a lists SNPs inside or within 2kb of named genes or very frequently observed putative transcripts that are as yet unnamed. Table S3b lists SNPs that are not within or near the named or frequently occuring genes. Many of these are non-synonymous SNPs in as yet unnamed genes. Tables S4a and S4b (Excel files with multiple sheets) list all the SNP locations that are plotted in Figs. 10,11,12,13,14. As compared with Table S3a, Table S4a contains the SNPs appearing in more than 20% of the classification test sets for the number of SNPs *N <* 40 or unnamed genes where the SNP appears in more than 80% of the test sets. Table S4b contains the SNPs appearing in more than 20% of the test sets but not within a named gene. These may be relevant but as yet uncharacterized genes, as in Table S3b

#### 6.4 Temporal classification of genomes from Malawi

Despite the general spatiotemporal inconsistencies in sampling rates, there is an opportunity to study the time dependence of genomic identity for parasites originating in Malawi. Figure18 show the results of a SSPOC analysis for four sets of ten repeated test and train model runs using the scrambled data and four sets of ten repetitions for the properly time-separated data. The genomic locations of the SSPOC SNPs for 2012-14 shows some known drug resistance loci such as *dhfr* and *dhps*, as well as the presence of SSPOC SNPs near *set1* as was previously found here for geographic separation. The scrambled data shows less concentration of SNPs at known loci. MAF values for the random permutation SSPOC SNPs are small, meaning that these SNPs are not very frequent in the population and might just be noise, while the SSPOC SNPs for temporal classification often have large MAF This genetic divergence with time is not as strong (the ratio is much closer to 1:1) as for the geographic separation, which is not surprising given that only two seasons have passed among the same population with assumedly similar selective pressures.

**Figure 7:**
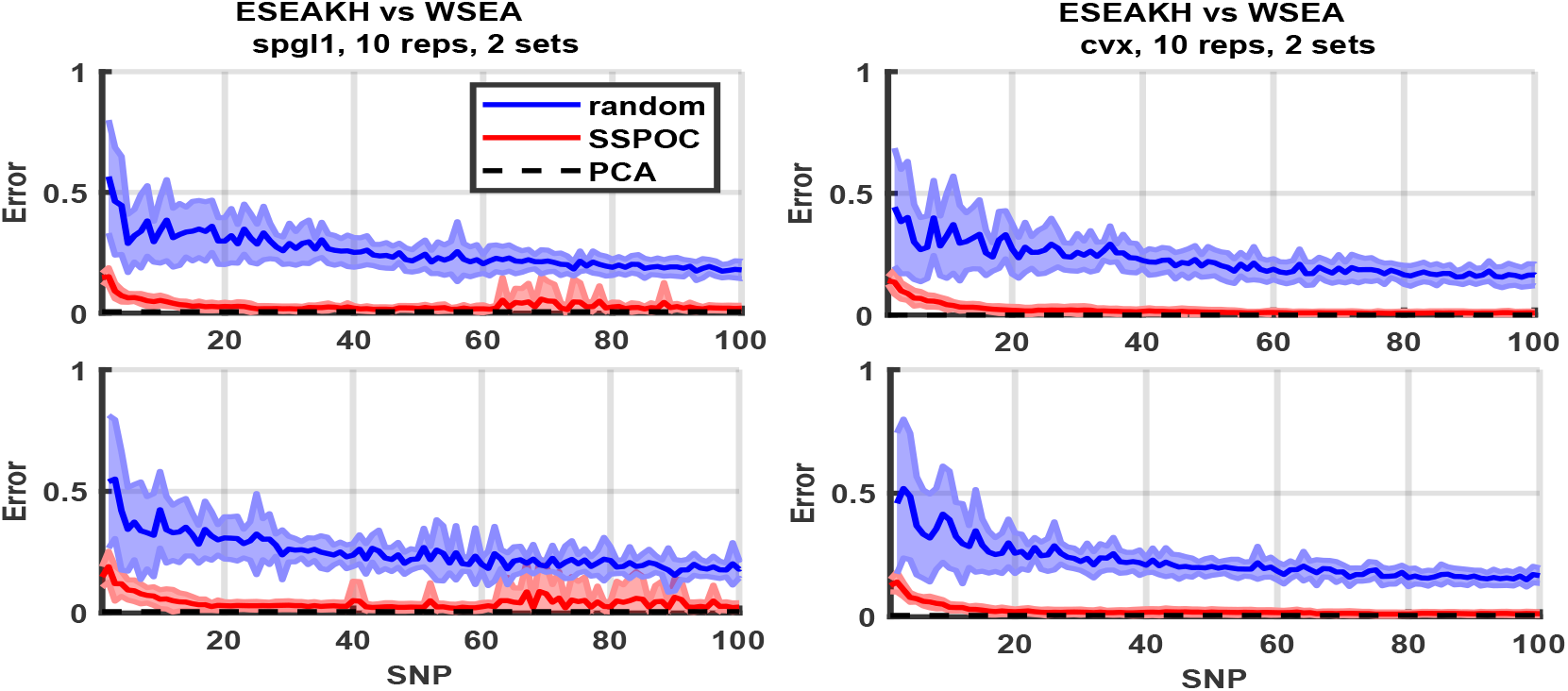
Comparison between numerical methods SPGL [61] and CVX [62, 63] for implementing the sparse optimization step in the sensor placement problem. Each method is compared with the high-fidelity principal component analysis (PCA) using 20 modes (black dashed lines near zero error). Both methods are compared to choosing randomly located SNP positions for a range of SNP set sizes up to 100 SNPs. While CVX seems to produce a more stable result for larger numbers of SNPs, SPGL is very accurate with respect to the optimal PCA result.

**Figure 8:**
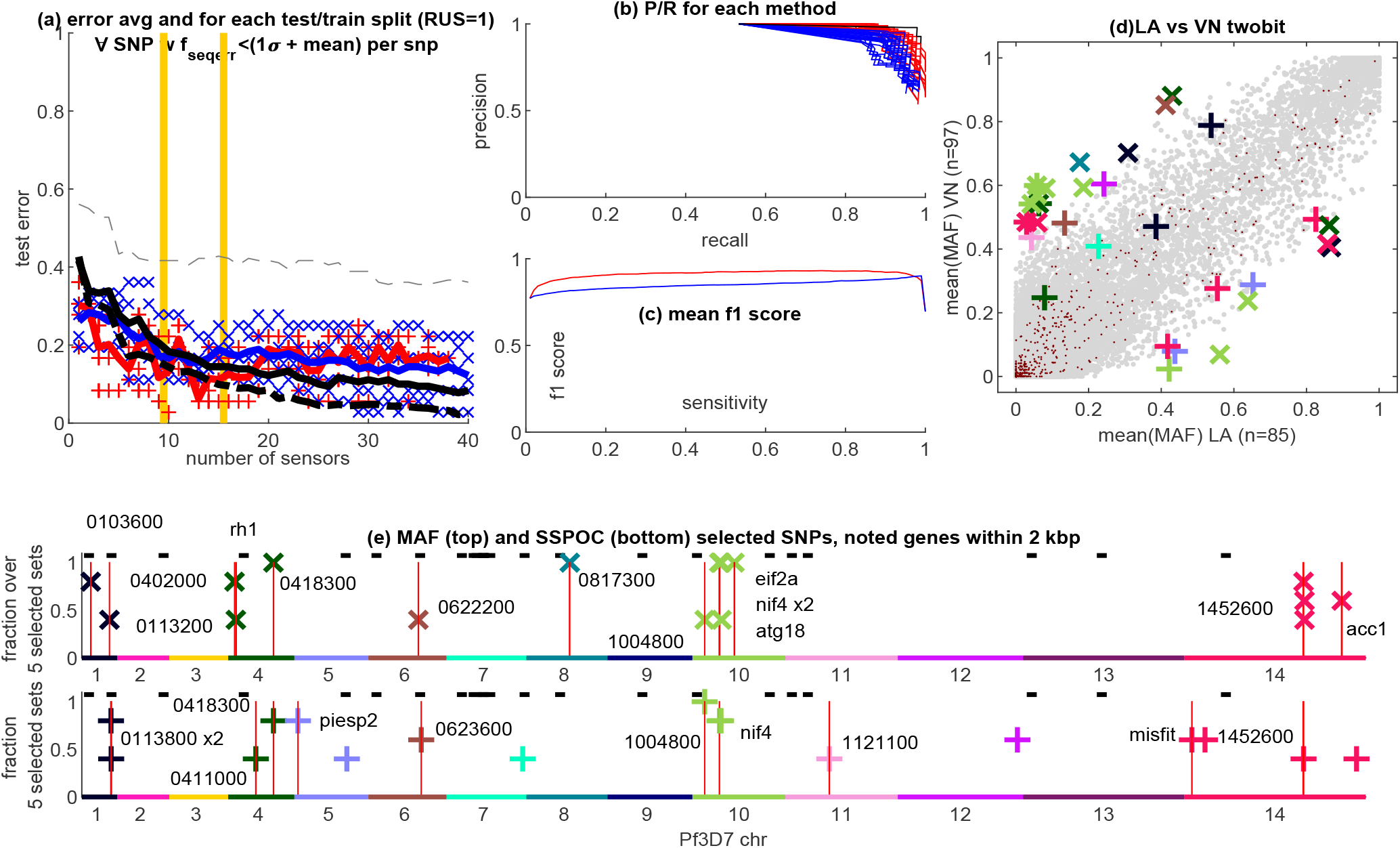
Additional panels for Laos v Vietnam to accompany Fig. 3. Panel a) shows the accuracy of classification for several methods. Red crosses represent SSPOC for each of the test/train splits used at each number of SNP sensors used to find the sparse solution. Blue x shows the same for MAF-ranking from 1 to 40 SNPs. The red trace is the average error over SSPOC test sets while the blue trace is the average for MAF-rank. The light dashed black line is the accuracy with a random set of SNPs (discrete sensors). The solid black line is the PCA accuracy for 1 to 40 modes using the test data set. The heavy dashed line is the PCA accuracy using the training data. Panel b) shows precision/recall for each subset of data, with SSPOC in red, MAF-rank in blue, and PCA on test data in black. Panel c) shows the average f1-score across all SSPOC and MAF-rank subsets. Panel d) shows the scatter plot for the sample-averaged MAF-rank compared between collection sites. Panel e) shows the location of SSPOC and MAF-rank SNPs across the genome with known genes labeled. The vertical axis in these genome plots shows the fraction of unique classification test sets (varying in both test/train split and number of SNPs allowed from 1 - 40) in which that SNP appeared.

**Figure 9:**
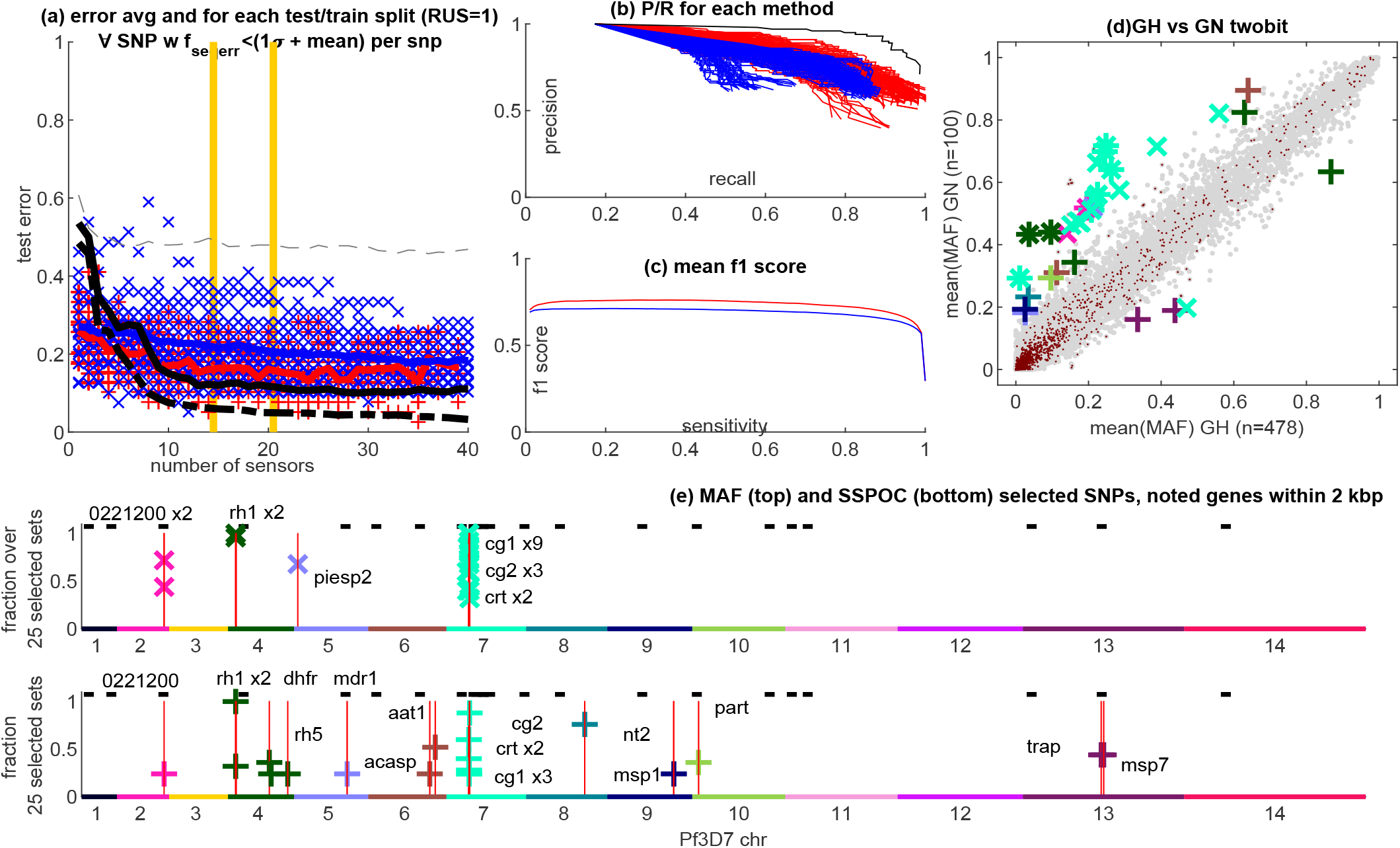
Additional panels for Guinea v Ghana to accompany Fig. 3. See Fig. 8 for description.

**Figure 10:**
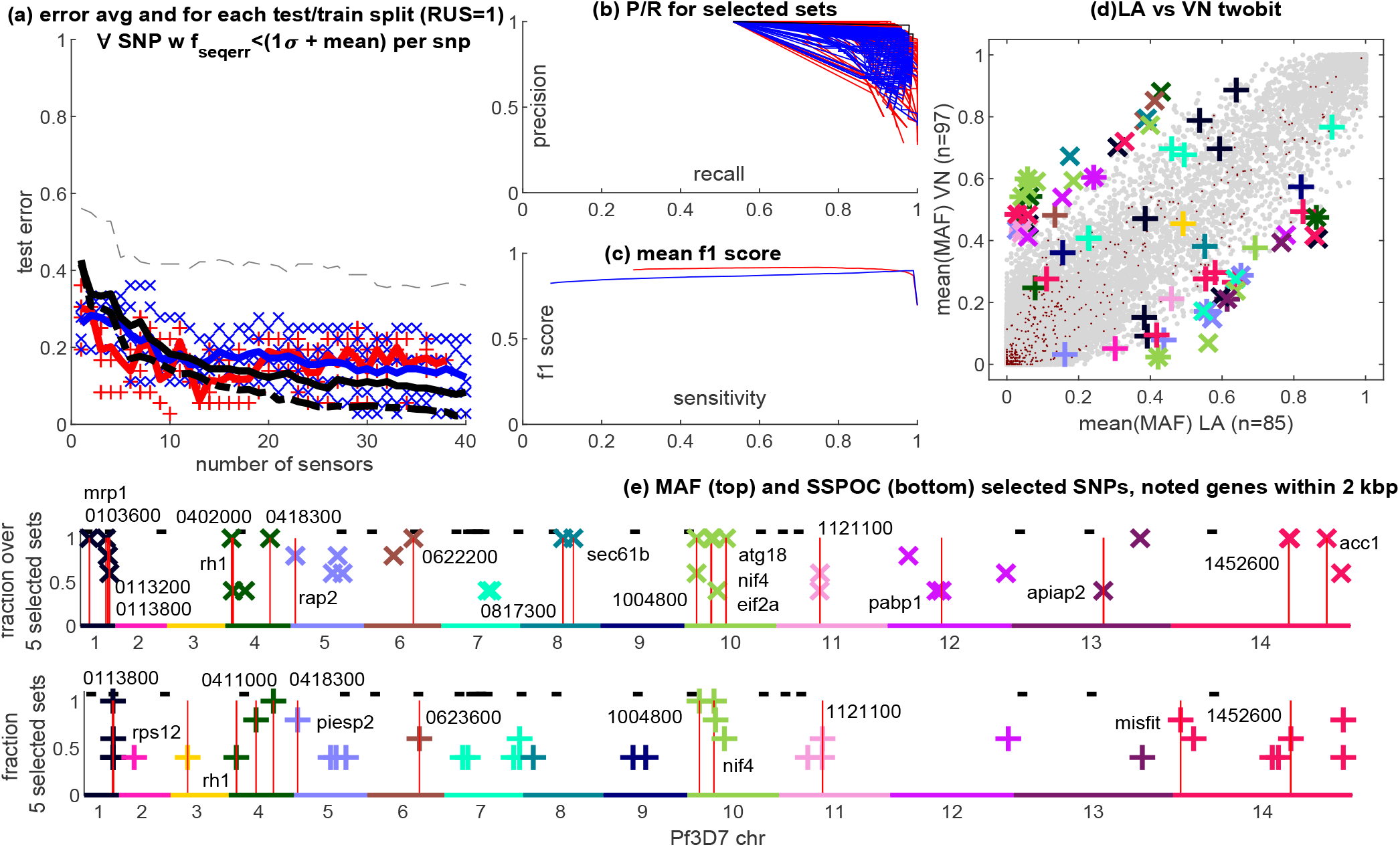
Full set of SNPs for Laos v Vietnam. Panel a) shows the accuracy of classification for several methods, where all other panels use all the sets of SNPs represented here. Red crosses represent SSPOC for each of the test/train splits used at each number of SNP sensors used to find the sparse solution. Blue x shows the same for MAF-ranking from 1 to 40 SNPs. The red trace is the average error over SSPOC test sets while the blue trace is the average for MAF-rank. The light dashed black line is the accuracy with a random set of SNPs (discrete sensors). The solid black line is the PCA accuracy for 1 to 40 modes using the test data set. The heavy dashed line is the PCA accuracy using the training data. Panel b) shows precision/recall for each subset of data, with SSPOC in red, MAF-rank in blue, and PCA on test data in black. Panel c) shows the average f1-score across all SSPOC and MAF-rank subsets. Panel d) shows the scatter plot for the sample-averaged MAF-rank compared between collection sites. Panel e) shows the location of SNPs from all sets derived from SSPOC and MAF-rank across the genome with known genes labeled. The vertical axis in these genome plots shows the fraction of unique classification test sets (varying in both test/train split and number of SNPs allowed from 1 - 40) in which that SNP appeared.

**Figure 11:**
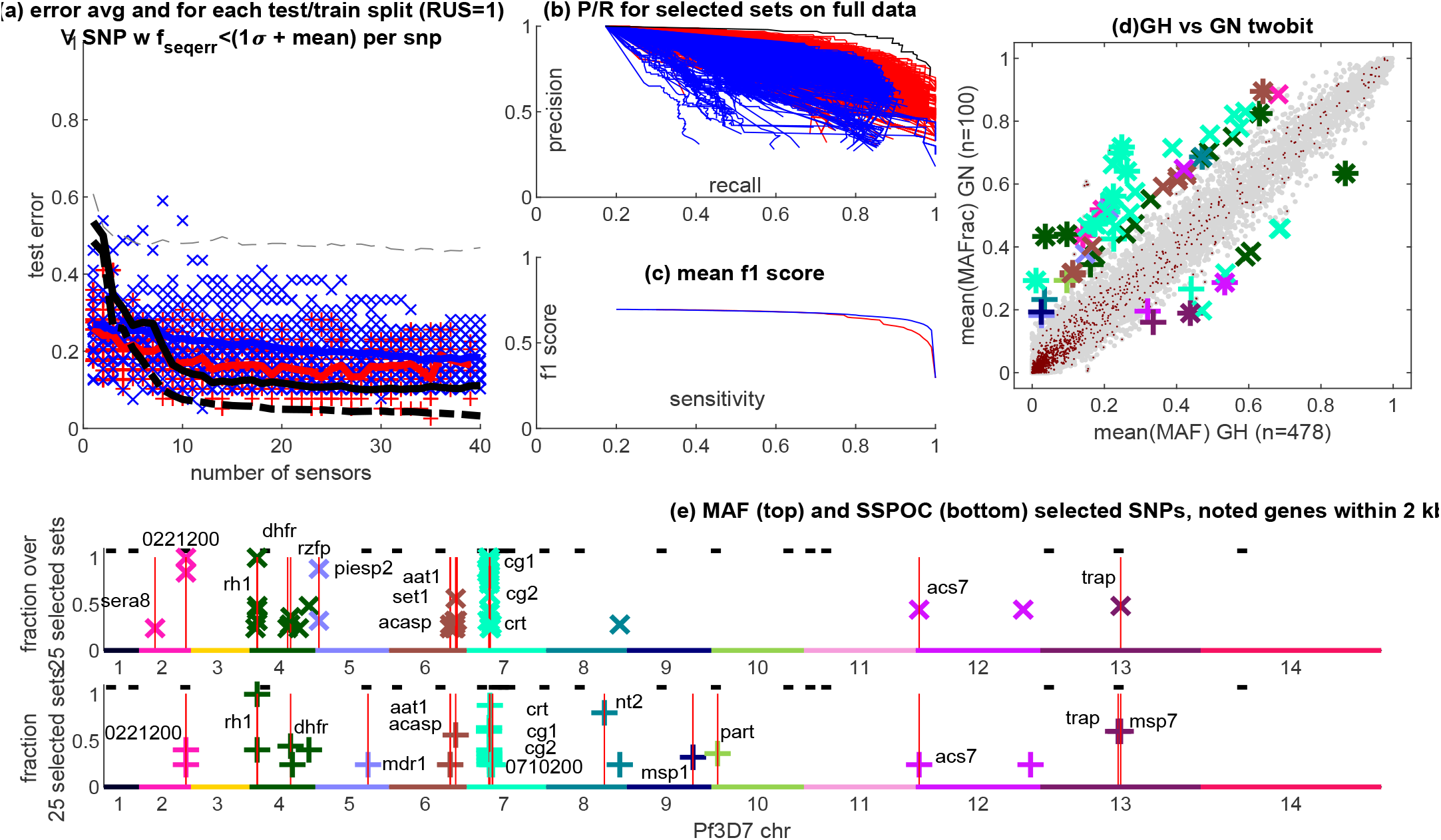
Full set of SNPs for Ghana v Guinea, see Fig. 10 for description.

**Figure 12:**
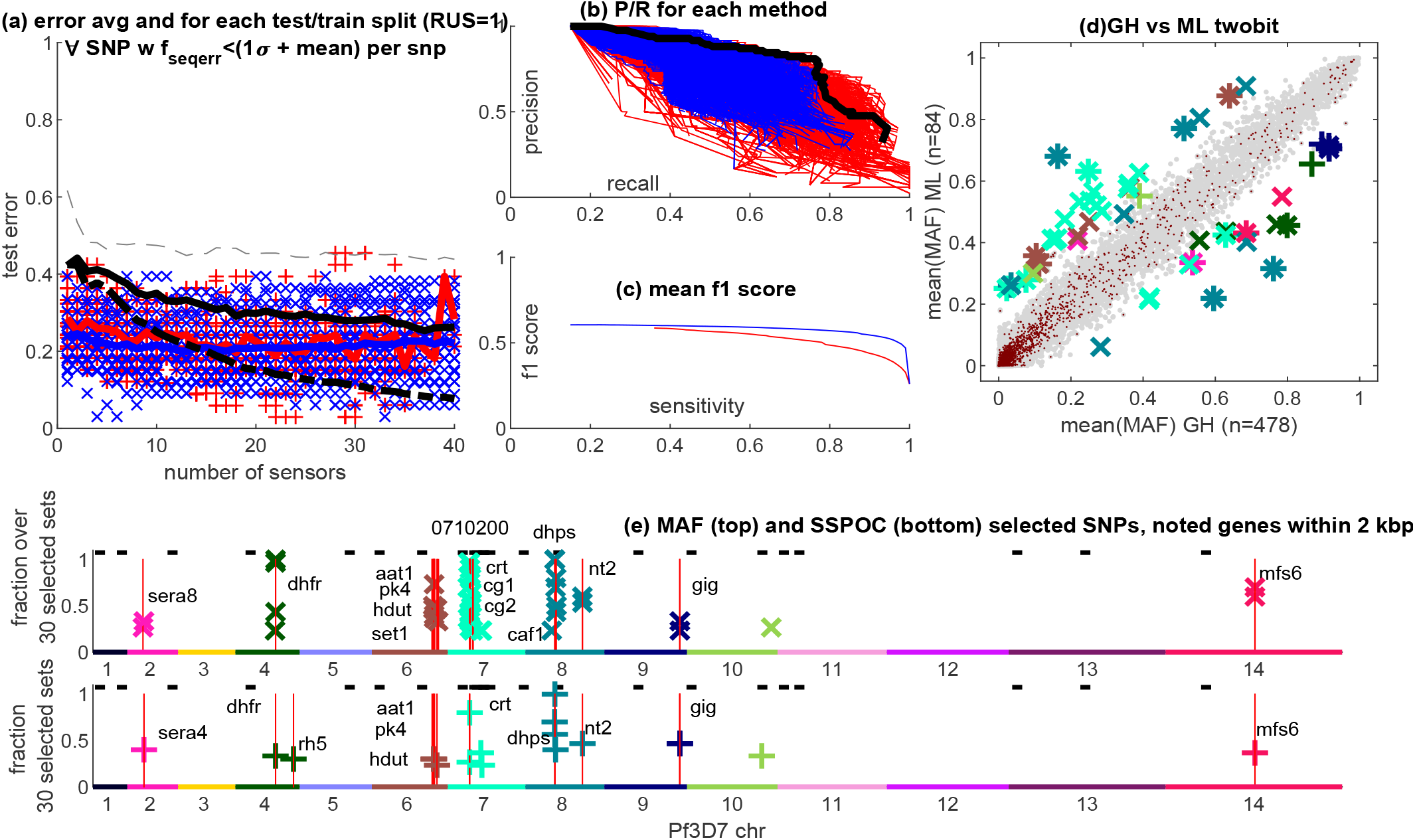
Full set of SNPs for Ghana v Mali, see Fig. 10 for description.

**Figure 13:**
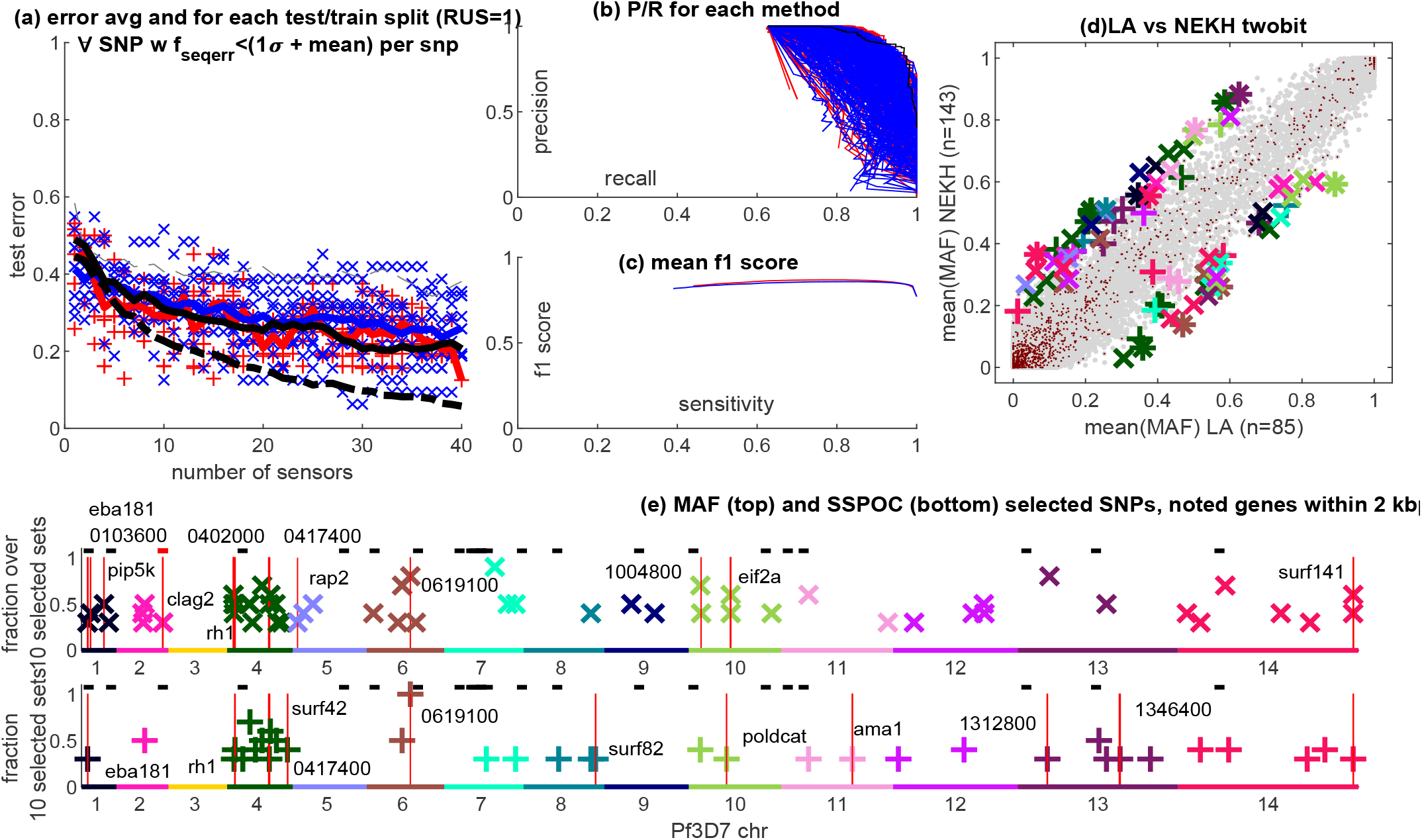
Full set of SNPs for Laos v NE Cambodia, see Fig. 10 for description.

**Figure 14:**
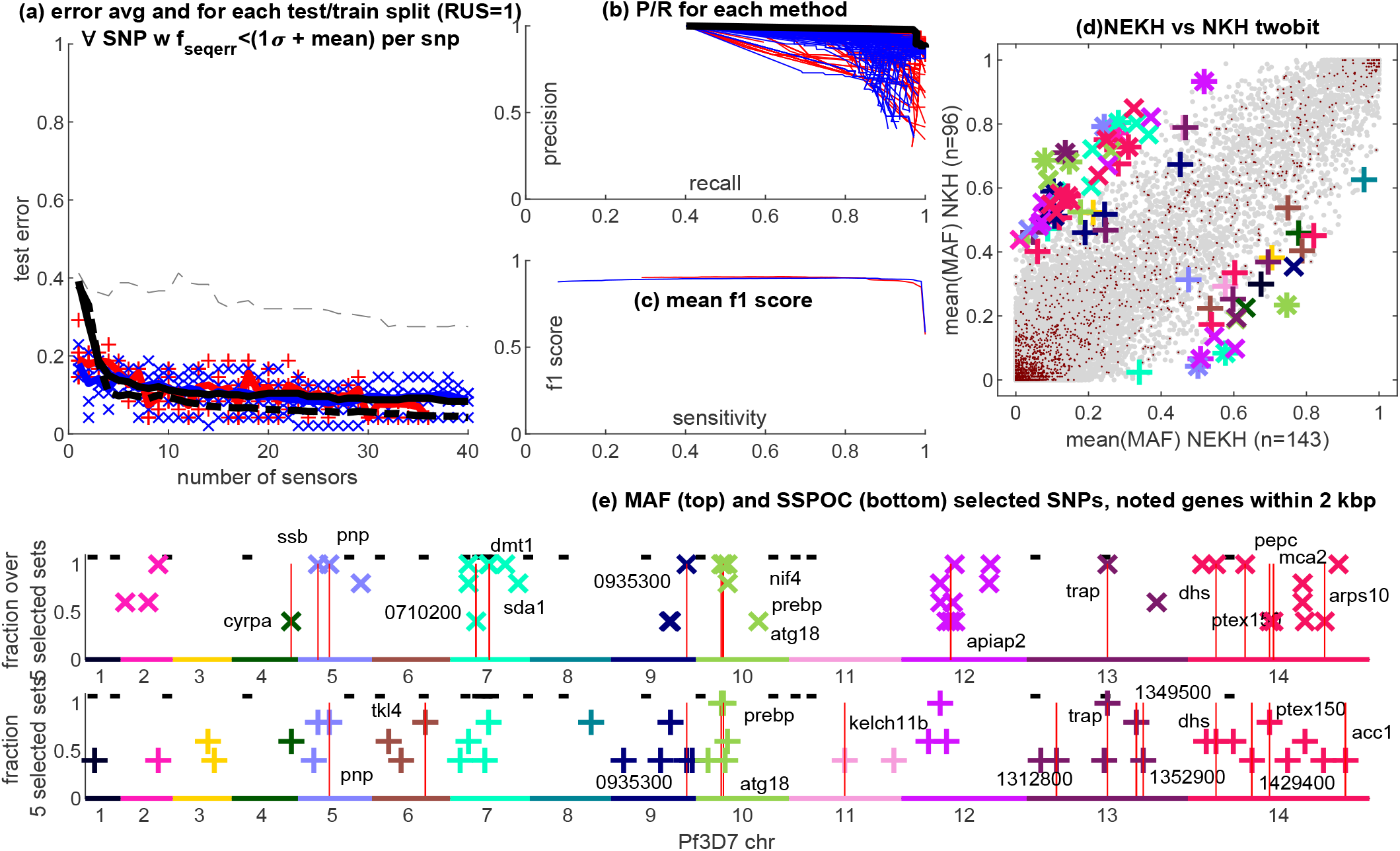
Full set of SNPs for N Cambodia v NE Cambodia, see Fig. 10 for description.

**Figure 15:**
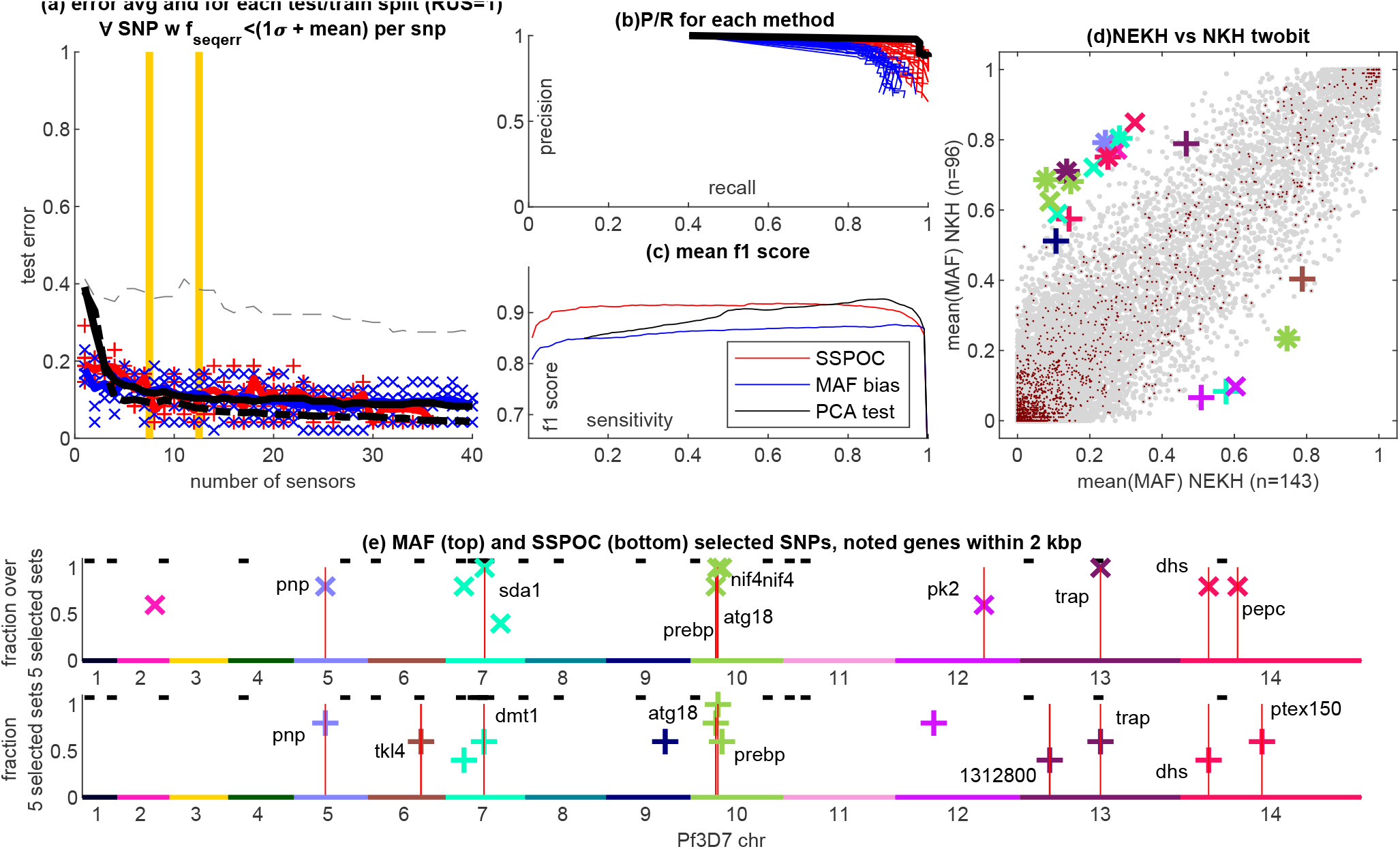
Reduced set of SNPs for N Cambodia v NE Cambodia, similar to Fig. 8 but with the mean f1 score for PCA on the test data shown. This mean value is taken over all test sets from 1 to 40 PCA modes.

**Figure 16:**
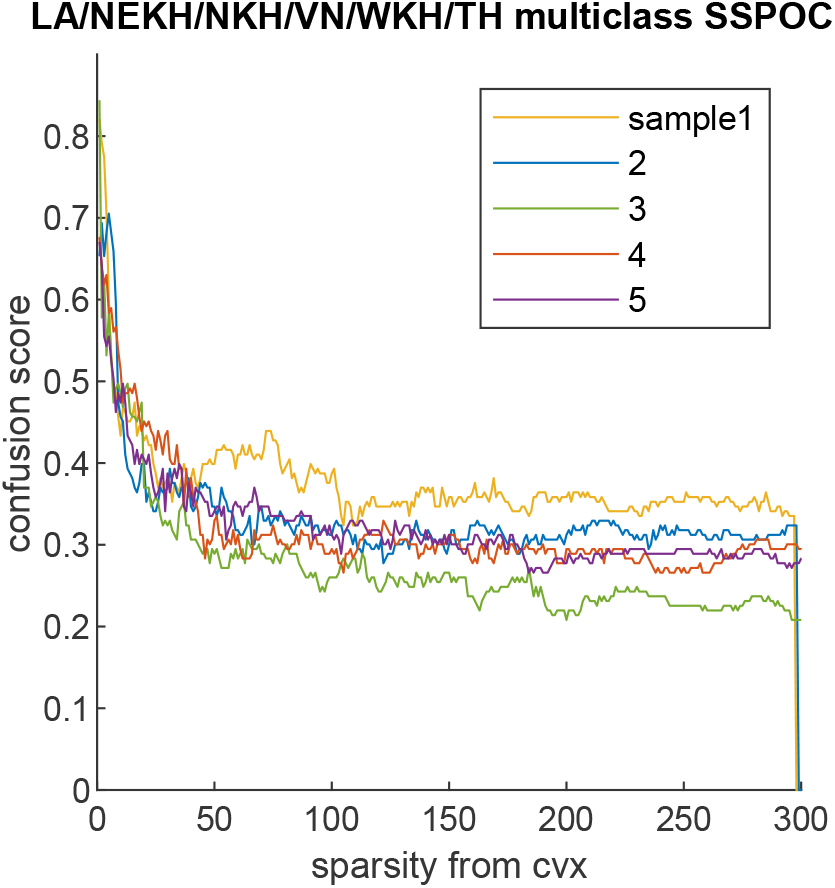
Trend in the confusion score for each of the five test/train splits used for the six-class SE Asia optimization shown in Fig. 4.

**Figure 17:**
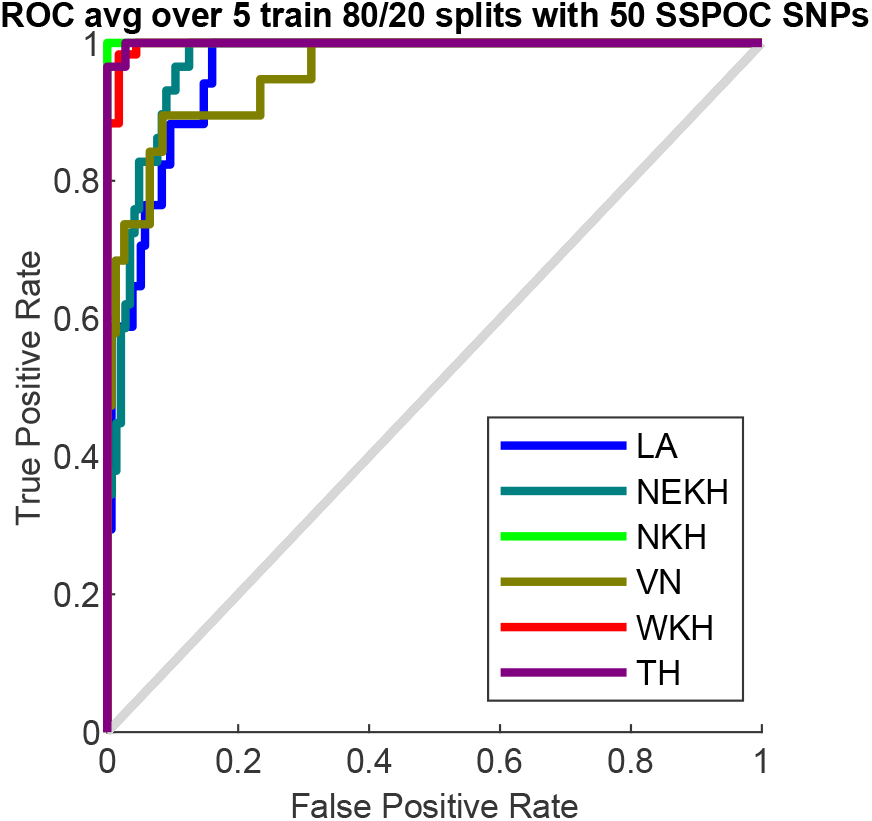
ROC curves for each of the classes averaged over five test/train splits used for the six-class SE Asia optimization shown in Fig. 4.

**Figure 18:**
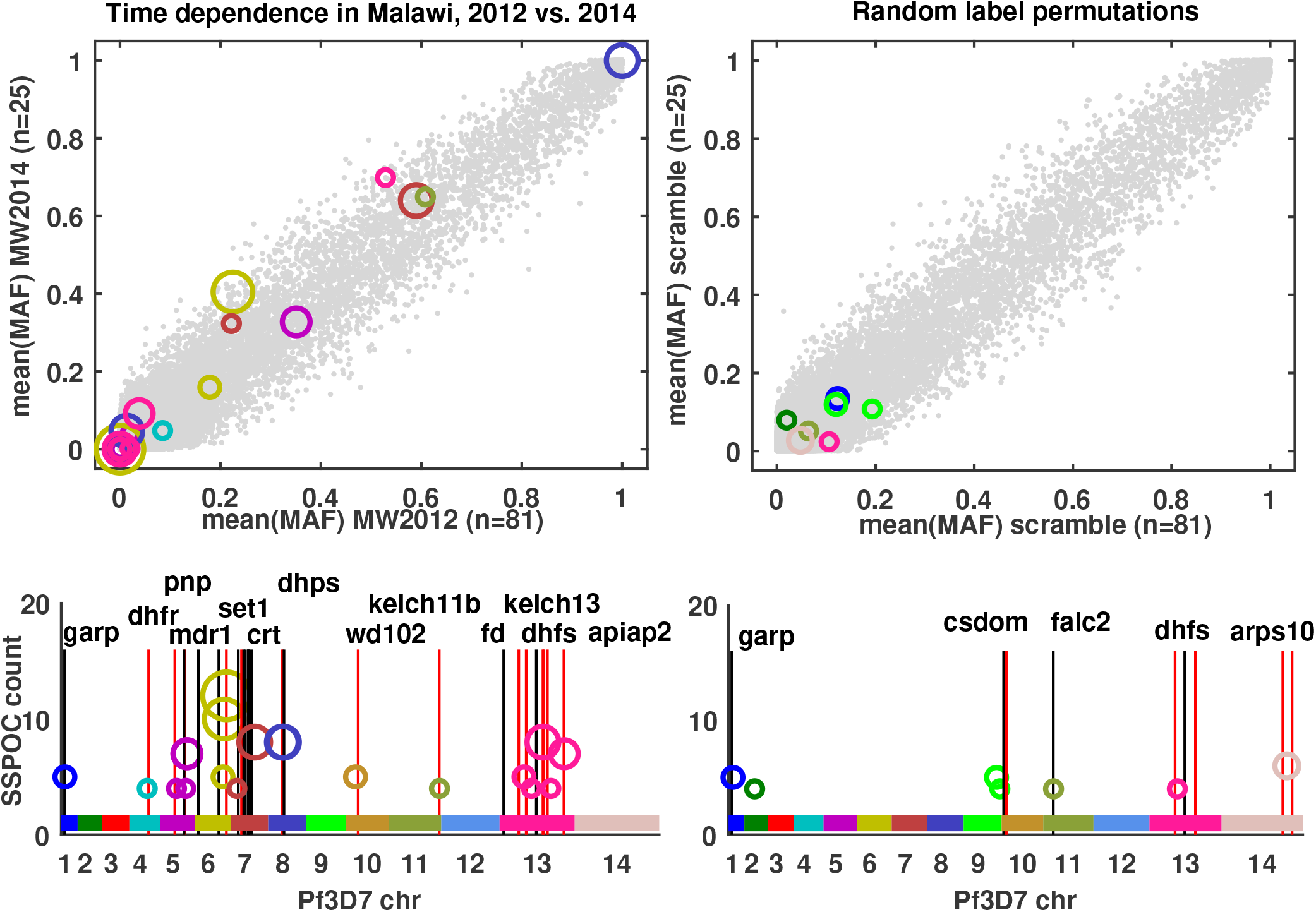
Minor allele frequency and genomic location of SSPOC SNPS most effective for separating years 2012 and 2014 for *P. falciparum* genomes. On the left, results using reported labels for genomes from Chikwawa, Malawi collection site in 2012 and 2014. On the right, results from SSPOC when randomizing the labels for the genomes. Note that only *N* = 25 samples were collected in 2014 compared to *N* = 81 samples in 2012.

## Notes

### Competing Interest Statement

The authors have declared no competing interest.

